# Causal mediation analysis of the neuroprotection of *APOE2* through lipid pathways

**DOI:** 10.1101/2025.01.03.25319984

**Authors:** Qingyan Xiang, Judith J. Lok, Nicole Roth, Stacy L. Andersen, Thomas T. Perls, Zeyuan Song, Anatoli I. Yashin, Jonas Mengel-From, Gary J. Patti, Paola Sebastiani

**Affiliations:** Department of Biostatistics, Vanderbilt University Medical Center, Nashville, TN, United States; Department of Mathematics and Statistics, Boston University, Boston, MA, United States; Section of Geriatrics, Department of Medicine, Boston University Chobanian & Avedisian School of Medicine, Boston, MA, United States; Institute for Clinical Research and Health Policy Studies, Tufts Medical Center, Boston, MA, United States; Biodemography of Aging Research Unit, Social Science Research Institute, Duke University, Durham, NC, United States; Department of Public Health, University of Southern Denmark, Denmark; Department of Genetics, Washington University in St. Louis, St. Louis, MO, United States; Department of Chemistry, Washington University in St. Louis, St. Louis, MO, United States; Department of Medicine, Washington University in St. Louis, St. Louis, MO, United States; Tufts University, School of Medicine, Boston, MA, United States; Data Intensive Study Center, Tufts University, Boston, MA

**Keywords:** Apolipoprotein E, Cognition, Longevity, Mediation analysis, Digital Assessment

## Abstract

**Background:** Recent studies have revealed a strong association between the e2 allele of the Apolipoprotein E (*APOE2)* gene and lipid metabolites. In addition, *APOE2* carriers appear to be protected from cognitive decline and Alzheimer’s disease. This correlation supports the hypothesis that lipids may mediate the protective effect of *APOE2* on cognitive function, thereby providing potential targets for therapeutic intervention.

**Methods:** We conducted a causal mediation analysis to estimate both the direct effect of *APOE2* and its indirect effect through 19 lipid species on cognitive function, using metrics from the digital Clock Drawing Test (CDT) in 1291 Long Life Family Study (LLFS) participants. The CDT metrics included think-time, ink-time, and their sum as total-time to complete the test.

**Results:** Compared to carriers of the common *APOE3*, *APOE2* carriers completed the CDT significantly faster. Two lipids showed protective mediation when elevated in the blood, resulting in shorter CDT think-time (CE 18:3), ink-time (TG 56:5), and total completion time (CE 18:3 and TG 56:5). Elevated TG 56:4, in contrast, showed deleterious mediation resulting in increased ink-time. The combined indirect effect through all lipids significantly mediated 23.1% of the total effect of *APOE*2 on total-time, reducing it by 0.92s (95% CI: 0.17, 2.00). Additionally, the sum of total indirect effect from all lipids also mediated 27.3% of the total effect on think-time, reducing it by 0.75s, and 13.6% of the total effect on ink-time, reducing it by 0.17s, though these reductions were statistically insignificant. Sensitivity analysis yielded consistent results of the combined indirect effects and total effects and identified additional significant lipid pathways (CE 22:6, TG 51:3, and TG 54:2).

**Conclusions:** We found that the combined indirect effect through all lipids could mediate 10%-27% of the total direct effect of *APOE2* on CDT times. We identified both protective and deleterious lipids, providing insights for new therapeutics targeting those lipids to modulate the protective effects of *APOE2* on cognition.

## Introduction

The apolipoprotein E (*APOE*) gene, a crucial gene in lipid metabolism, has been extensively studied for its association with cognitive function and late-onset Alzheimer’s Disease (AD) [1–4]. The *APOE* gene has three well-characterized alleles—e2, e3, and e4 —that are defined by combinations of the Single Nucleotide Polymorphisms (SNPs) rs7412 and rs429358. Among these alleles, the e3 allele is the most common in Non-Hispanic and White individuals and is considered neutral. The e4 allele is considered a major genetic determinant for AD risk and cognitive decline [5–7], and the e2 allele is associated with increased human longevity [8] and decreased risk for AD and cognitive decline [9–13].

Extensive research has focused on the direct effect of *APOE* alleles on the risk for AD and cognitive decline. However, there have been limited investigations on the effect of *APOE* that is explained or mediated through molecular pathways [14–16]. *APOE* plays an important role in lipid metabolism, a modifiable risk factor for cognitive decline [17], and different *APOE* alleles are characterized by replicated lipid profiles [14, 18, 19]. Therefore, characterizing and quantifying the role of lipids as mediators can improve our understandings of the mechanism of *APOE* on cognition, providing insights for new therapeutics targeting these lipid pathways.

In this exploratory study, our objective is to investigate the effect of *APOE2* on cognitive function, as assessed by the Clock Drawing Test (CDT), and to examine whether this effect is mediated through lipid metabolites. The CDT is a widely used screening tool for global cognitive dysfunction, with results shown to correlate with cognitive performance. Specifically, faster CDT completion time have been associated with improved processing speed and logical memory [20]. Using results from a digitally administered CDT, we computed two metrics: think-time, which refers to the time spent holding the pen without drawing, and ink-time, which refers to the time spent drawing the features of the clock on the paper. We also computed the sum of think-time and ink-time as the total-time to complete the test. Focusing on a list of lipids previously identified as significantly associated with *APOE2* [21], we performed a causal mediation analysis to estimate (1) the direct effect of *APOE2*, (2) the indirect effects of *APOE2* through these lipids, and (3) the total effect of *APOE2* on CDT think, ink, and total-time among 1291 Long Life Family Study (LLFS) participants [18].

## Methods

### Participants

#### Long Life Family Study (LLFS)

The LLFS is a multicenter, multigeneration study that enrolled 4,953 family members from 539 families who exhibit healthy aging and longevity. Participants were first enrolled between 2006 and 2009 at three American field centers (in Boston, Pittsburgh, and New York) and a Danish field center. The second in-person visit was completed during 2014-2017 for participants using the same protocols. Further details on the LLFS study can be found in reference [22]. All participants provided informed consent through their local Institutional Review Board, and the genetic and phenotypic data generated through 2017 are available through dbGaP (dbGaP Study Accession: phs000397.v1.p1). New data generated after 2017 will be distributed through the ELITE portal: https://eliteportal.synapse.org/Explore/Projects/DetailsPage?shortName=LLFS.

The CDT, a common screening test for global cognitive dysfunction and for a range of neurological and psychiatric illnesses [23], was added to the neuropsychological assessment protocol at the second in-person assessment. The CDT was administered using a digital pen that recorded spatial-temporal features of the test performance. These features were extracted by the THink software developed by Massachusetts Institute of Technology and Lahey Hospital and Medical Center [24], which generates think-time (time spent holding the pen without drawing), ink-time (time spent drawing the features of the clock on the paper), and their sum as total-time to complete the test. We focused on these three measures as outcomes of the causal mediation analysis.

### *APOE* genotype data

*APOE* alleles were determined from genotypes of two SNPs, rs7412 and rs429358, that were generated using Whole Genome Sequencing [22]. The e2 allele was defined by the combination rs7412=T and rs429358=T; the e3 allele was defined by the combination rs7412=C and rs429358=T; and the e4 allele by rs7412=C and rs429358=C. This study focused on the comparison between genotype group *APOE3* versus genotype group *APOE2* in LLFS participants, where the genotype groups were defined as: *APOE3*=e3e3 (reference group) and *APOE2*=e2e2 or e2e3. Carriers of one or more copies of the e4 alleles were excluded.

### Lipids

For lipid measurements, this study used blood collected during the first in-person visit between 2006 and 2009. Lipids were analyzed from plasma by liquid chromatography/mass spectrometry (LC/MS) as described previously [21, 25]. In brief, lipids were first isolated by using solid-phase extraction kits. Lipids were then separated by reversed-phase chromatography prior to being measured on an Agilent 6545 quadrupole time-of-flight mass spectrometer. Samples were analyzed in batches of approximately 90. Pooled samples, reference materials, and internal standards were used for quality control and batch correction, thereby ensuring high data quality. A detailed description of these methods was described in a prior report [26].

In the analysis, we started from 24 lipids that were associated with *APOE2* at 5% FDR in Sebastiani et. al, (2024) [24]. These include sterol lipids (CEs), sphingolipids (DGs), glycerolipids (TGs), dHexCer_NS 34:1, and dHexCer_NS 41:1, and they were log-transformed and standardized before the mediation analysis.

### Statistical analysis

To study how the effect of *APOE* on cognition is mediated by lipids, we performed a causal mediation analysis to estimate the direct and indirect effects of *APOE2* on CDT performance. The primary components of the causal mediation analysis included these elements:

- <u>Exposure</u>: *APOE2* genotype group *versus APOE3* genotype group (reference group).
- <u>Mediators</u>: Lipids level data that were log-transformed and then standardized.
- <u>Confounders measured at baseline</u>: age at enrollment, sex, education, lipid-lowering medication usage, and indicator of young/old generation based on whether the birth year > 1935. Please see Supplementary Figure 1 for the distribution of birth year in LLFS participants.
- <u>Outcomes</u>: (1) CDT total-time derived by summing (1.a) CDT think-time and (1.b) CDT ink-time.

**Figure 1:**
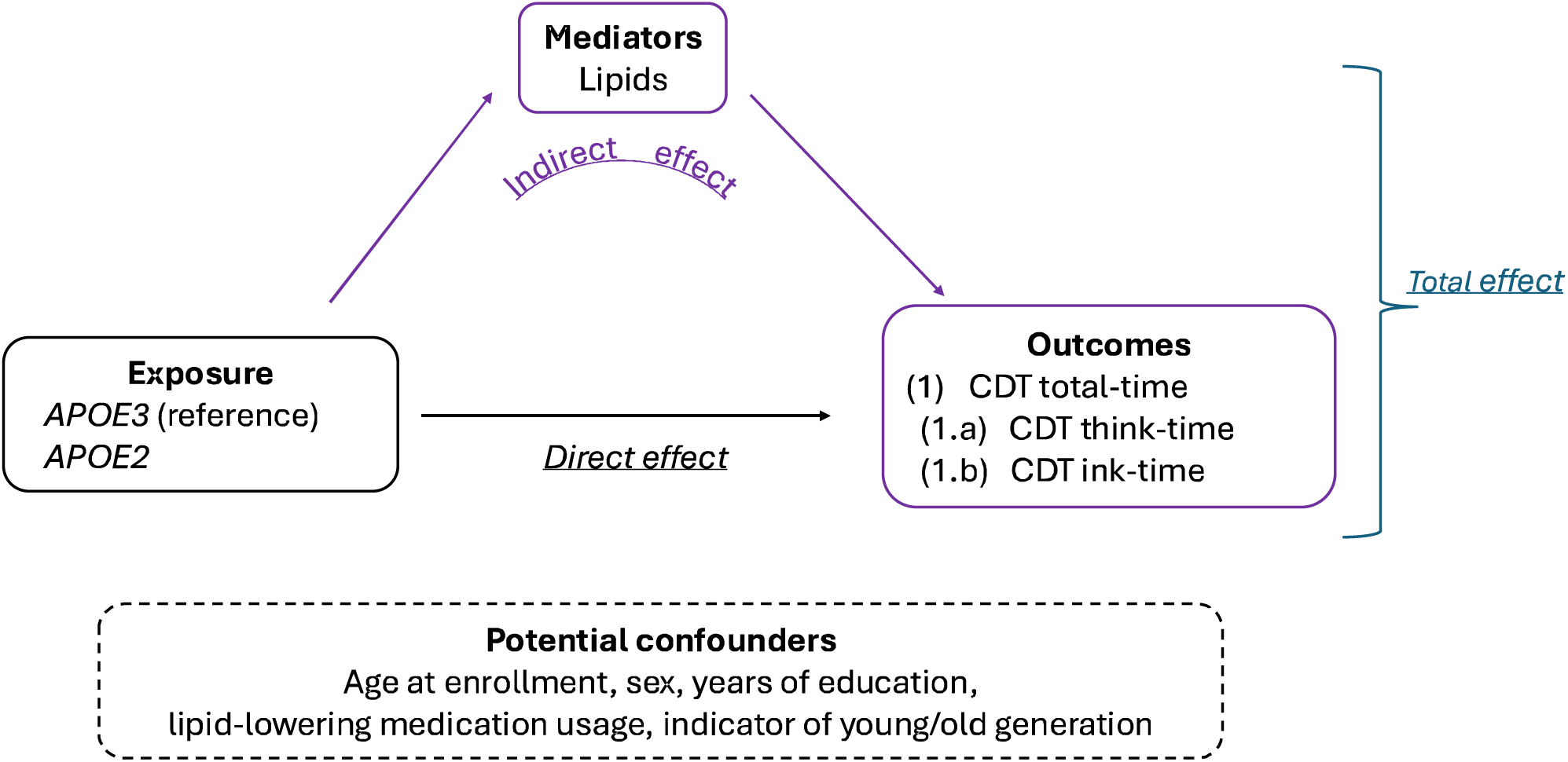
Components of the mediation analysis in this study: exposure, mediators, potential confounders, and outcomes. CDT: Clock Drawing Test.

In this analysis, the lipids (mediators) are measured at the first visit and the CDT results (outcomes) are measured at the second visit, satisfying the temporal ordering assumption of the mediation analysis, where the causal sequences follow exposure -> mediator -> outcome. In addition, to avoid multicollinearity of lipids in the regression models, we excluded highly correlated lipids. Specifically, we excluded one lipid species from each pair with a Pearson correlation greater than 0.85.

We used a regression-based approach for the causal mediation analysis, as outlined in references [27, 28]. First, we fit the *mediator regression model* for each of the lipids, where the independent variable was the *APOE* genotype group, adjusted for all the confounders. Next, we fit the *outcome regression model* for each of the three outcomes listed above, and the independent variables were the *APOE* genotype group and all lipids, adjusted for all the confounders. Then, we combined the estimates from these regression models [27] to estimate the direct effect and indirect effect of *APOE2* on the CDT times (See details in the supplementary file). To account for within-family correlations, we used generalized estimating equations to estimate standard errors in all regression models with an exchangeable covariance matrix based on family IDs.

In addition, we performed a sensitivity analysis by applying a backward stepwise variable selection in the outcome regression model, based on the AIC criterion. We then repeated the entire causal mediation analysis only including the lipids retained in the regression models after variable selection. In reporting the mediation analysis results, we present:

(1) the <u>direct effect</u> of *APOE2* on the CDT times.
(2) The <u>individual indirect effect</u> mediated through each lipid, where *APOE*2 affects each lipid, which in turn affects the CDT time.
(3) the <u>combined indirect effect</u>, which is the sum of all indirect effects via lipid pathways.
(4) the <u>total effect</u>, which is the sum of the direct effect and the combined indirect effect.

We generated 95% confidence intervals for these effect estimates using the bootstrap with Efron’s percentile method [29] with 1000 replicates. We used R 4.1.3 for all analyses and all scripts are available from <u>QM&DS Tufts Medical Center (github.com)</u>.

## Results

### Participant and lipid characteristics

Our analysis included 1291 participants with *APOE* genotype data and plasma lipids measured at the first visit and CDT data from the second visit. Table 1 summarizes the characteristics of the LLFS participants included in this analysis. Among those participants, *APOE3* carriers and *APOE2* carriers had similar ages at enrollment, proportion of females, and years of education. However, *APOE3* carriers included a larger proportion of participants taking lipid lowering medications (33%) than *APOE2* carriers (19%).

**Table 1:** Characteristics of participants who were included in the analysis. Continuous variables are summarized with median and interquartile range. Discrete variables are summarized with count and percentage. *APOE3*=e3e3, *APOE2*=e2e2 or e2e3. CDT: Clock Drawing Test.

| Characteristic | Overall, N = 1291 | *APOE3*, N = 1040 | *APOE2*, N = 251 |
| --- | --- | --- | --- |
| Age at enrollment | 62.0 (56.5, 70.0) | 62.0 (57.0, 70.0) | 62.0 (56.0, 70.5) |
| Age at visit two | 72.0 (65.0, 79.0) | 72.0 (65.0, 79.0) | 71.0 (65.0, 80.0) |
| Sex |  |  |  |
| Female | 739 (57%) | 604 (58%) | 135 (54%) |
| Male | 552 (43%) | 436 (42%) | 116 (46%) |
| Years of education | 14.0 (10.0, 15.0) | 14.0 (10.0, 15.0) | 14.0 (10.0, 14.0) |
| Lipid-lowering medication usage | 386 (30%) | 338 (33%) | 48 (19%) |
| CDT total-time | 32.3 (25.9, 41.6) | 32.8 (26.2, 41.9) | 30.2 (24.8, 40.5) |
| Ink-time | 13.5 (10.8, 16.8) | 13.7 (11.0, 16.9) | 12.8 (9.9, 16.1) |
| Think-time | 18.5 (14.0, 25.4) | 18.6 (14.0, 25.7) | 17.9 (13.8, 24.3) |

As described in the method section, we excluded lipids that were highly correlated (see details in the supplementary file). Table 2 lists the 19 lipid species that were included in the analysis. These lipids belong to the super classes of sterol lipids, sphingolipids, and glycerolipids.

**Table 2:** List of 19 lipid species included in the analysis.

| **Standardized Name (RefMet)** | **Super class** |
| --- | --- |
| **CE 18:2** | Sterol Lipids |
| **CE 18:3** | Sterol Lipids |
| **CE 20:4** | Sterol Lipids |
| **CE 22:5** | Sterol Lipids |
| **CE 22:6** | Sterol Lipids |
| **Cer 33:1** | Sphingolipids |
| **DG 38:5** | Glycerolipids |
| **dHexCer_NS 34:1** | NA |
| **dHexCer_NS 41:1** | NA |
| **TG 51:0** | Glycerolipids |
| **TG 51:3** | Glycerolipids |
| **TG 53:1** | Glycerolipids |
| **TG 54:2** | Glycerolipids |
| **TG 56:1** | Glycerolipids |
| **TG 56:3** | Glycerolipids |
| **TG 56:4** | Glycerolipids |
| **TG 56:5** | Glycerolipids |
| **TG 58:3** | Glycerolipids |
| **TG 58:6** | Glycerolipids |

### Primary analysis of digital CDT

#### Mediator regression

Table 3 shows the results of the mediator regression that describes the associations between *APOE2* and lipids (log scale and standardized). Consistent with previous work [18], the estimated associations between *APOE* and lipids in the LLFS participants included in this analysis were predominantly statistically significant. Compared to *APOE3*, *APOE2* carriers had lower levels of sterol lipids (CE) and higher levels of glycerolipids (TG), with the exception of TG 51:3 that was 19.7% lower (1-exp(−0.22)=19.7%, p<0.01).

**Table 3:**
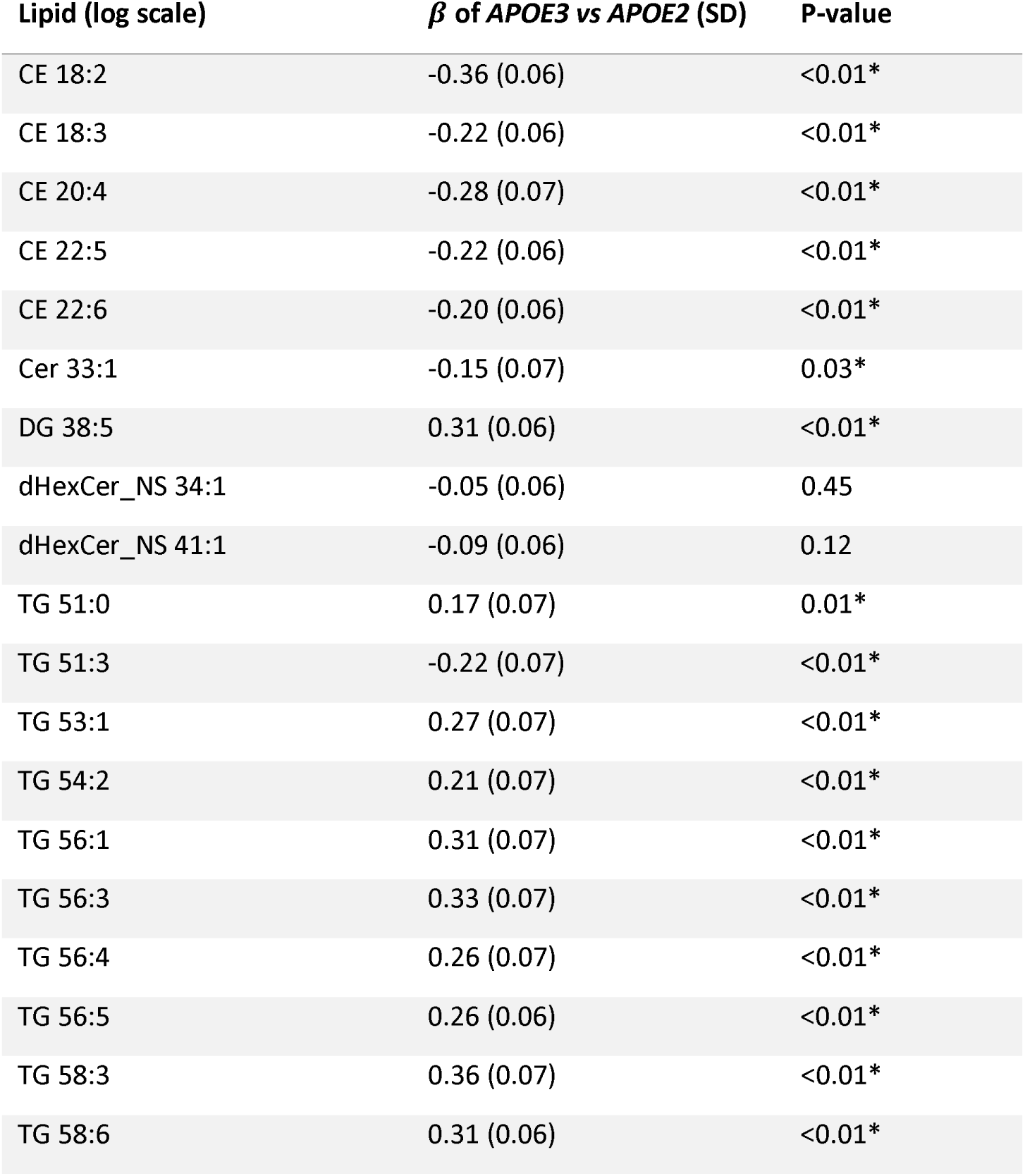
Results of the mediator regression in the primary analysis. Dependent variable: lipids (log scale and standardized). Independent variables: *APOE3* (reference group) vs *APOE2*, age at enrollment, sex, education, lipid-lowering medication usage, and indicator for young/old generation. *β*: estimated coefficients of *APOE3* (reference) versus *APOE2* on each lipid. SD: standard deviation. *: p-value that reaches the significance level of 0.05.

#### Outcome regression

Table 4 shows the results of the outcome regressions of CDT total-time, think-time, and ink-time. The results show that *APOE2* had a statistically significant negative association with total-, think-, and ink-time, after adjusting for all lipids, age, sex, education and other confounders. Among all the lipids, CE 18:3 had a significantly positive association with total-time (*β* = 1.50, *p* = 0.03) and think-time (*β* = 1.27, *p* = 0.01, TG 56:4 showed a significantly positive association with ink-time (*β* = 1.04, *p* = 0.02), and TG 56:5 showed a significantly negative association with total-time (*β* = −4.48, *p* = 0.03) and think-time (*β* = −1.58, *p* < 0.01). No other lipid species was significantly associated with any of the CDT times.

**Table 4:** Results of the outcome regression in the primary analysis of digital CDT times. Dependent variable: Digital CDT total-time, ink-time, and think-time. Independent variables: *APOE3* (reference group) vs *APOE2*, lipids (log scale and standardized), age at enrollment, sex, education, lipid-lowering medication usage, and indicator of young/old generation. *β*: estimated coefficients. SD: standard deviation. Bold font indicates the estimated coefficients reaching the significance level of 0.05.

|  | Total-time |  | Think-time |  | Ink-time |  |
| --- | --- | --- | --- | --- | --- | --- |
| | $\beta$ (SD) | p-value | $\beta$ (SD) | p-value | $\beta$ (SD) | p-value |
| *APOE2* | **-3.07 (0.98)** | **<0.01** | **-1.98 (0.72)** | **0.01** | **-1.06 (0.39)** | **<0.01** |
| CE 18:2 | -0.60 (0.87) | 0.49 | -0.19 (0.66) | 0.77 | -0.42 (0.31) | 0.17 |
| CE 18:3 | **1.50 (0.67)** | **0.03** | **1.27 (0.49)** | **0.01** | 0.24 (0.25) | 0.34 |
| CE 20:4 | 1.45 (1.03) | 0.16 | 0.86 (0.79) | 0.28 | 0.59 (0.34) | 0.09 |
| CE 22:5 | -1.32 (0.9) | 0.14 | -1.15 (0.69) | 0.09 | -0.19 (0.33) | 0.56 |
| CE 22:6 | 0.99 (0.83) | 0.23 | 1.1 (0.63) | 0.08 | -0.09 (0.29) | 0.76 |
| Cer 33:1 | 1.01 (0.65) | 0.12 | 0.77 (0.52) | 0.14 | 0.23 (0.2) | 0.25 |
| DG 38:5 | -1.85 (1.08) | 0.09 | -1.19 (0.77) | 0.12 | -0.65 (0.38) | 0.09 |
| dHexCer_NS 34:1 | 0 (0.78) | 1.00 | -0.03 (0.59) | 0.96 | 0.03 (0.27) | 0.92 |
| dHexCer_NS 41:1 | -0.85 (0.84) | 0.32 | -0.74 (0.6) | 0.22 | -0.09 (0.32) | 0.78 |
| TG 51:0 | -0.88 (1.46) | 0.55 | -0.43 (1.01) | 0.67 | -0.45 (0.61) | 0.46 |
| TG 51:3 | -1.48 (0.84) | 0.08 | -0.92 (0.62) | 0.14 | -0.57 (0.3) | 0.06 |
| TG 53:1 | 1 (1.92) | 0.60 | 0.48 (1.38) | 0.73 | 0.52 (0.75) | 0.49 |
| TG 54:2 | 2.3 (1.37) | 0.09 | 1.9 (1.01) | 0.06 | 0.4 (0.48) | 0.41 |
| TG 56:1 | -1.43 (1.41) | 0.31 | -0.93 (0.97) | 0.34 | -0.5 (0.55) | 0.37 |
| TG 56:3 | 0.27 (1.48) | 0.86 | 0.79 (1.08) | 0.47 | -0.48 (0.53) | 0.37 |
| TG 56:4 | 1.6 (1.22) | 0.19 | 0.57 (0.91) | 0.53 | **1.04 (0.46)** | **0.02** |
| TG 56:5 | **-4.48 (2.03)** | **0.03** | -2.93 (1.64) | 0.07 | **-1.58 (0.55)** | **<0.01** |
| TG 58:3 | 0.19 (1.19) | 0.87 | -0.23 (0.89) | 0.80 | 0.39 (0.39) | 0.31 |
| TG 58:6 | 0.75 (0.96) | 0.43 | 0.46 (0.74) | 0.54 | 0.31 (0.33) | 0.34 |

#### Indirect effect of APOE2 on CDT times through lipids

Figure 2 (A) shows the significant direct effect, as well as the significant lipid-mediated pathways for the indirect effects, and Table 5 and Figure 3 show all indirect effects of *APOE2* on the CDT times through each lipid pathway. Three lipid species appeared to significantly mediate the *APOE2* effects: CE 18:3 and TG 56:5 with a protective effect, and TG 56:4 with a deleterious effect (only on ink-time). The indirect effect of *APOE2* through CE 18:3 and TG 56:5 significantly reduced CDT total-time by 0.30 seconds (CE 18:3, 95% CI: −0.67, −0.03) and by 1.28 seconds (TG 56:5, 95% CI: −3.11, −0.15), respectively. In addition, the indirect effect of *APOE2* through CE 18:3 significantly reduced think-time by 0.25 seconds (CE 18:3, 95% CI: −0.54, −0.02), and through TG 56:5 significantly reduced ink-time by 0.45 seconds (95% CI: −0.98, −0.10). On the other hand, the indirect effect of *APOE2* through TG 56:4 significantly increased ink-time by 0.30 seconds (95% CI: 0.03, 0.69). Such effect through TG 56:4 on total-time was similarly deleterious but did not reach statistical significance (95% CI: −0.26, 1.35).

**Figure 2:**
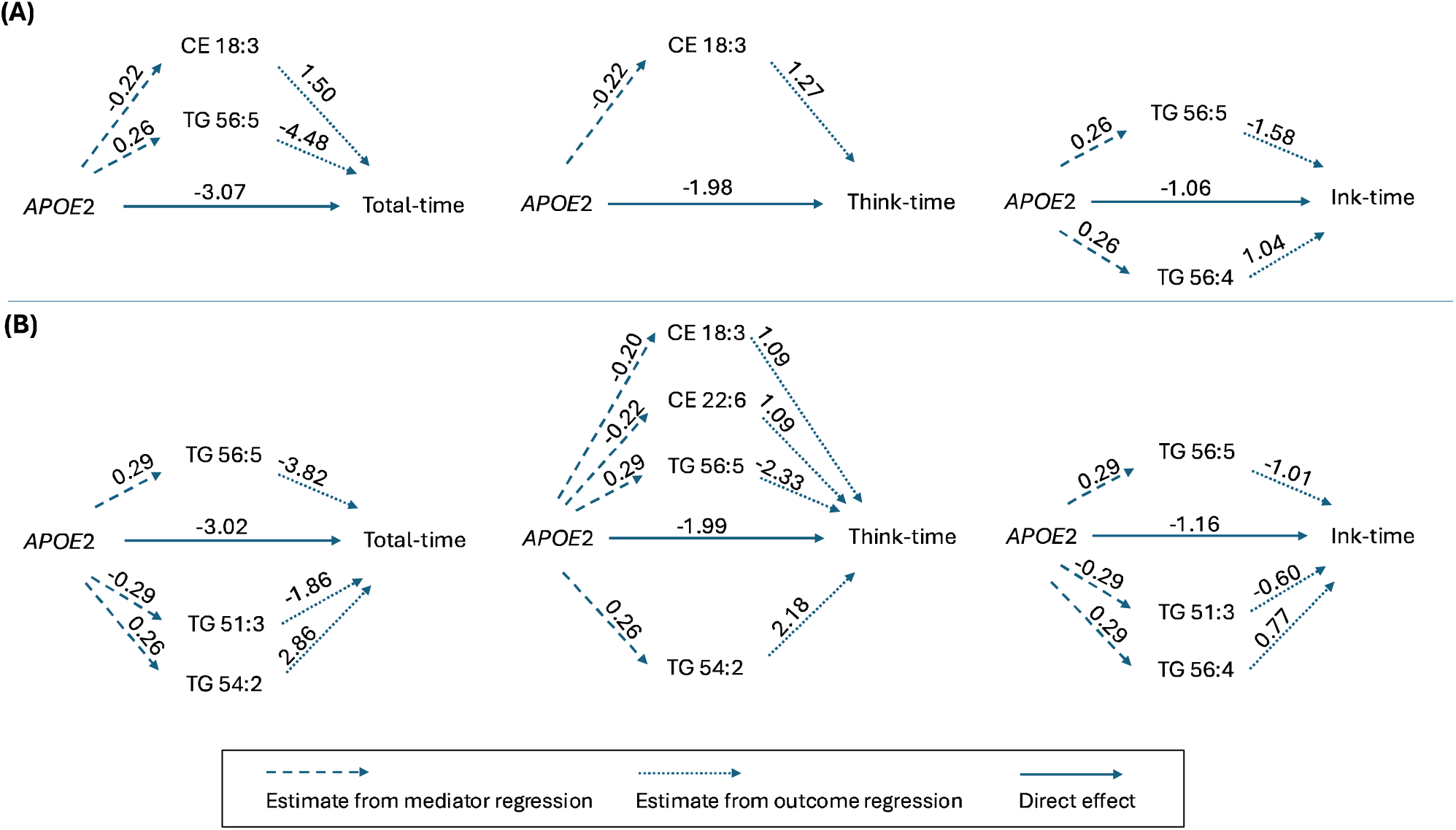
Mediation analysis results: significant direct and indirect effects in both primary analysis (Panel A) and sensitivity analysis (Panel B). The numbers on the left-side dashed lines represent the estimated associations between *APOE2* and the lipids. The numbers on the right-side dotted lines represent the estimated associations between the lipids and the CDT times. The dashed lines and dotted lines combined together represent the pathway of the indirect effect. The solid arrow represents the direct effect of *APOE2* on CDT times. Lipids with protective effect are above the solid arrow in the middle, e.g., CE 18:3; Lipids with deleterious effect are below the solid arrow in the middle, e.g., TG 56:4.

**Table 5:** Results of estimated indirect effects of *APOE2* on CDT times through each lipid pathway. 95% confidence intervals (CI) are generated using bootstrap. Protective lipids mediated the effect of *APOE2* to reduce CDT times, while deleterious lipids mediated the effect of *APOE2* to increase CDT times. Bold font indicates the effect estimates reaching the significance level of 0.05.

| Indirect effect estimates (95% CI) | Total time | Think time | Ink time |
| --- | --- | --- | --- |
| CE 18:2 | 0.22 (-0.52, 1.07) | 0.07 (-0.62, 0.65) | 0.16 (-0.06, 0.43) |
| CE 18:3 | **-0.30 (-0.67, -0.03)** | **-0.25 (-0.54, -0.02)** | -0.05 (-0.17, 0.06) |
| CE 20:4 | -0.43 (-1.17, 0.20) | -0.25 (-0.85, 0.26) | -0.17 (-0.43, 0.05) |
| CE 22:5 | 0.33 (-0.13, 1.01) | 0.29 (-0.09, 0.79) | 0.05 (-0.16, 0.23) |
| CE 22:6 | -0.22 (-0.7, 0.15) | -0.25 (-0.63, 0.03) | 0.02 (-0.11, 0.18) |
| Cer 33:1 | -0.17 (-0.55, 0.07) | -0.13 (-0.44, 0.07) | -0.04 (-0.15, 0.05) |
| DG 38:5 | -0.68 (-1.66, 0.22) | -0.44 (-1.08, 0.14) | -0.24 (-0.57, 0.07) |
| dHexCer_NS 34:1 | 0 (-0.17, 0.15) | 0 (-0.13, 0.12) | 0 (-0.06, 0.04) |
| dHexCer_NS 41:1 | 0.08 (-0.12, 0.44) | 0.07 (-0.08, 0.35) | 0.01 (-0.08, 0.13) |
| TG 51:0 | -0.18 (-1.01, 0.50) | -0.09 (-0.66, 0.38) | -0.09 (-0.46, 0.15) |
| TG 51:3 | 0.43 (-0.08, 1.09) | 0.27 (-0.12, 0.75) | 0.16 (-0.03, 0.42) |
| TG 53:1 | 0.31 (-1.05, 1.87) | 0.15 (-0.88, 1.16) | 0.16 (-0.29, 0.74) |
| TG 54:2 | 0.59 (-0.21, 1.66) | 0.49 (-0.13, 1.23) | 0.1 (-0.17, 0.42) |
| TG 56:1 | -0.55 (-1.91, 0.44) | -0.35 (-1.28, 0.47) | -0.19 (-0.67, 0.23) |
| TG 56:3 | 0.1 (-1.15, 1.32) | 0.29 (-0.6, 1.32) | -0.17 (-0.66, 0.29) |
| TG 56:4 | 0.47 (-0.26, 1.35) | 0.17 (-0.42, 0.80) | **0.3 (0.03, 0.69)** |
| TG 56:5 | **-1.28 (-3.11, -0.15)** | -0.84 (-2.27, 0.07) | **-0.45 (-0.98, -0.10)** |
| TG 58:3 | 0.08 (-1.03, 1.25) | -0.1 (-0.90, 0.68) | 0.17 (-0.17, 0.57) |
| TG 58:6 | 0.26 (-0.39, 1.14) | 0.16 (-0.45, 0.86) | 0.11 (-0.13, 0.38) |

**Figure 3:**
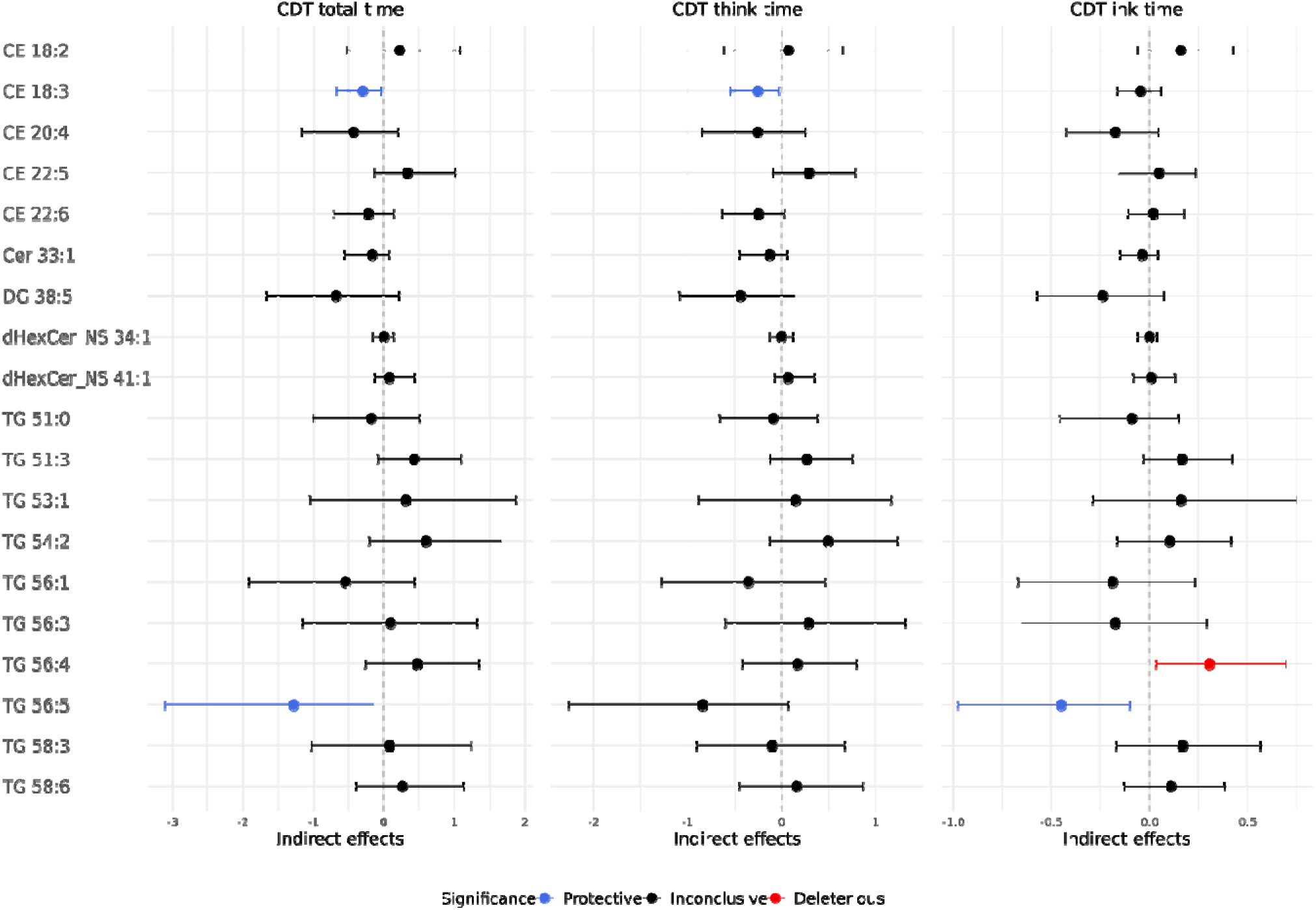
Primary analysis of indirect effects of *APOE2* on CDT times through each lipid pathway. Protective lipids mediated the effect of *APOE2* to reduce CDT times, while deleterious lipids mediated the effect of *APOE2* to increase CDT times.

#### Total and combined effects of APOE2 on CDT times

Table 6 summarizes all the results of the mediation analysis. Compared to *APOE*3, *APOE*2 carriers completed the CDT test 3.99 seconds faster (estimate: −3.99; 95% CI: −6.35, −1.23). This reduced time can be decomposed into a direct effect of 3.07 seconds (estimate: −3.07; 95% CI: −5.13, −1.06) and a significant combined indirect effect mediated through all lipids of 0.92 seconds (estimate: −0.92; 95% CI: −2.00, −0.17; mediated proportion: 23.1% of the total effect). Considering the two components of total-time, first, the think-time of *APOE2* carriers was 2.73 faster than *APOE3* carriers (estimate: −2.73; 95% CI: −4.66, −0.69). This reduced time can be decomposed into a direct effect of 1.98 seconds (estimate: −1.98; 95% CI: - 3.57, −0.31) and a combined indirect effect mediated through all lipids of 0.75 seconds (estimate: - 0.75; 95% CI: −1.92, 0.36; mediated proportion: 27.3% of the total effect). Second, the ink-time of *APOE2* carriers was 1.23 seconds faster than *APOE3* carriers (estimate: −1.23; 95% CI: −2.13, −0.30). This reduced time can be decomposed into a direct effect of 1.06 seconds (estimate: −1.06; 95% CI: - 1.89, −0.30) and a combined indirect effect of 0.17 seconds (estimate: −0.17; 95% CI: −0.58, 0.26; mediated proportion: 13.6% of the total effect).

**Table 6:**
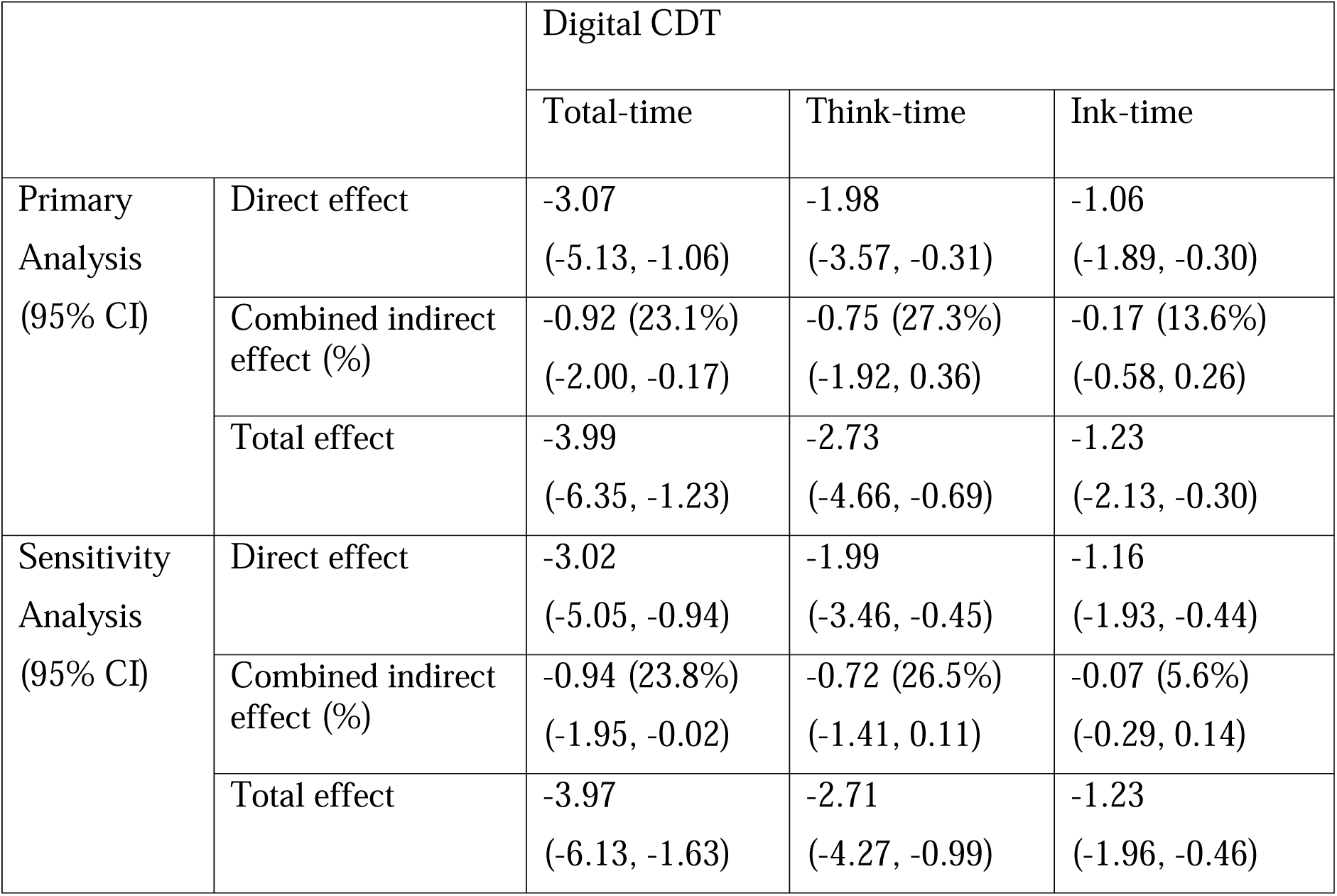
Summary of mediation analyses of *APOE2* on the digital CDT times with 95% CI. These estimates are negative because the *APOE2* group has a shorter CDT completion time compared to *APOE3*. Direct effect: the effect of *APOE2* on the CDT time that do not involve lipids. Combined indirect effect: the sum of all indirect effects via lipid pathways, the percentage indicating the proportion of the total effect mediated. Total effect: the sum of the direct effect and the combined indirect effect. CDT: Clock Drawing Test. CI: Confidence Interval.

### Sensitivity analysis

For each outcome, we conducted a sensitivity analysis by performing a variable selection of the outcome regression model, and we repeated the entire causal mediation analysis using the lipids that remained in the final selected model. Results of the mediator regression and the outcome regression in the sensitivity analyses for these CDT times are shown in Supplementary Tables 1-2.

For this sensitivity analysis, Figure 3 and Supplementary Table 3 show the individual indirect effect on the CDT times through each lipid pathway, and Figure 4 (B) shows the significant direct effect and indirect effects of *APOE2* on the CDT times. Consistently, TG 56:5 and CE 18:3 remained as protective mediators, and TG 56:4 remained as a deleterious mediator for ink-time. In addition, three new lipids were also found to significantly meditate the *APOE* effects: CE 22:6 with a protective effect, and TG 51:3 and TG 54:2 with a deleterious effect. The indirect effect of *APOE2* through CE 22:6 significantly reduced think-time by 0.24 seconds (estimate: −0.24; 95% CI: −0.50, 0). The indirect effect of *APOE2* through TG 51:3 increased think-time by 0.54 seconds (95% CI: 0.07, 1.27), and through TG 54:2 increased both total-time and think-time by 0.74 seconds (95% CI: 0.22, 1.47) and by 0.56 seconds (95% CI: 0.17, 1.21), respectively.

**Figure 4:**
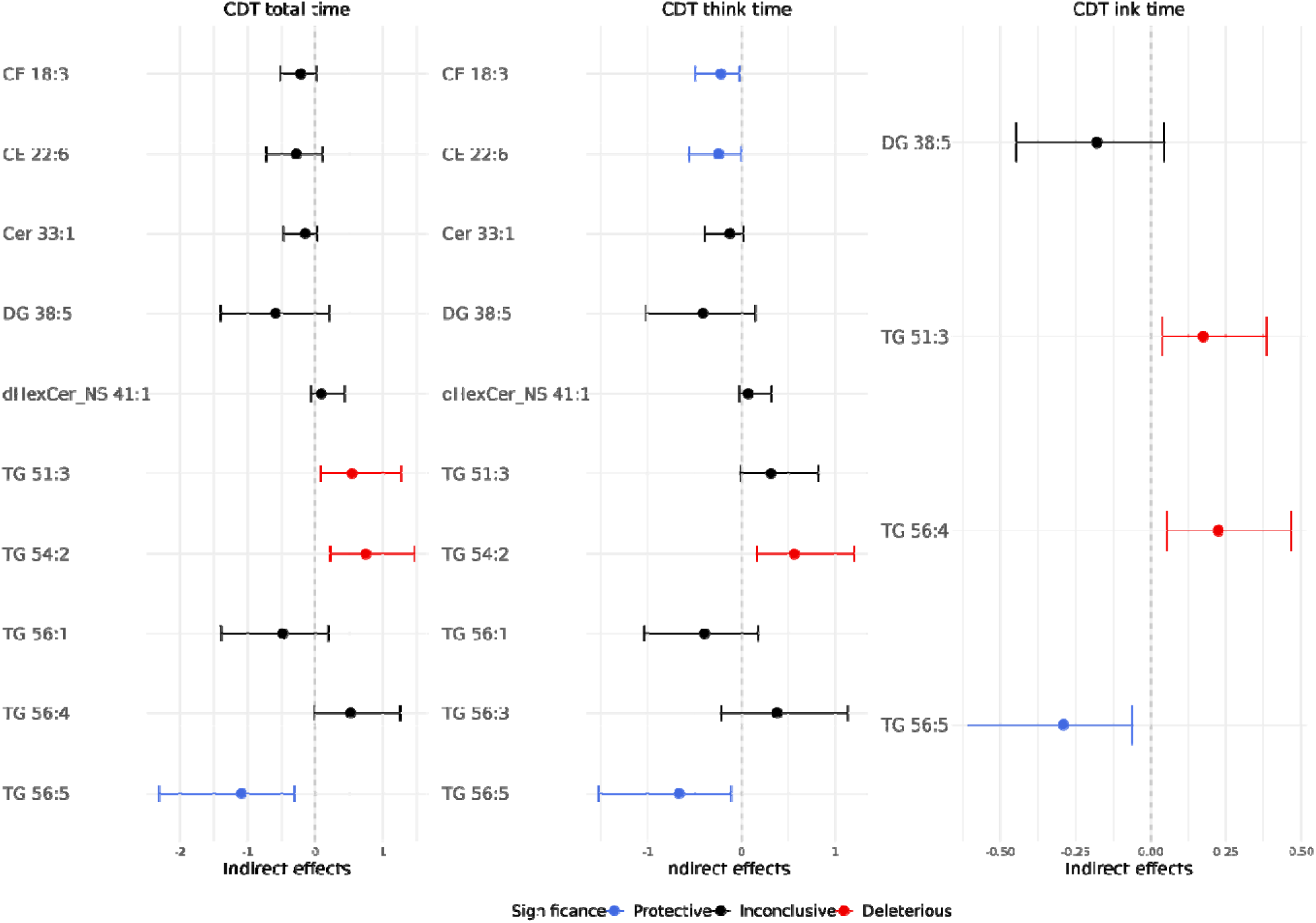
Sensitivity analysis of indirect effects of *APOE2* on CDT times through each lipid pathway. Stepwise variable selection with AIC criterion was applied in the sensitivity analysis, and hence each CDT time had different lipids remained in the mediation analysis.

Table 3 also summarizes the results of the mediation analysis of this sensitivity analysis for CDT times. The total effect of *APOE2* on each outcome in the sensitivity analysis was almost the same compared to the primary analysis. For total-time, the combined indirect effect remained statistically significant, mediating the effect of *APOE2* for 0.94 reduced seconds (95% CI: −1.95, −0.02; mediated proportion: 23.8%). Additionally, for both total-time and think-time, the combined indirect effect contributed to a similar mediated proportion to the primary analysis. However, for ink-time, the mediated proportion of the combined indirect effect decreased from 13.6% to 5.6% compared to the primary analysis, possibly due to fewer lipids remaining in the model after variable selection.

## Discussion

### Overview

We performed an analysis to investigate the potential mediating role of lipids on the effect of *APOE2* on CDT total-time, think-time, and ink-time. In the primary analysis, we identified a significant protective direct effect of *APOE2* on CDT total-time, think-time, and ink-time, significant protective indirect effects through two elevated lipids (CE 18:3 and TG 56:5), and a significant deleterious indirect effect through one lipid (TG 56:4). Overall, the combined indirect effect through all lipid pathways investigated here significantly mediated 23.1% of the total effect of *APOE2* on CDT total-time for a 0.92s faster completion time compared to *APOE3*. The sensitivity analysis revealed a significant protective effect through one additional lipid (CE 22:6) and a significant deleterious effect through two additional lipids (TG 51:3 and TG 54:2). Compared to the primary analysis, the total effect on all CDT times in the sensitivity analysis were almost the same, and the combined indirect effect on total-time and think-time contributed to a similar mediated proportion of the total effect.

### Discussion

Although the role of *APOE* in aging and cognition has been extensively studied [1], the mechanisms by which the *APOE2* allele protects against cognitive decline and promotes longevity remain elusive [30]. The strong correlations between *APOE2* alleles and many lipid species suggest that lipids in the blood may mediate the genetic effect of *APOE* on cognitive function [14, 15, 25], where individuals with certain lipid profile can present “reduced/increased” risk of cognitive decline. Our findings in the primary and sensitivity analyses identified six lipids (CE 18:3, CE 22:6, TG 51:3, TG 54:2, TG 56:4, TG 56:5) that significantly mediated the effect of *APOE2*, highlighting their potential as therapeutic targets for preserving cognitive function during aging. Specifically, our analyses identified two mechanisms that could be therapeutic targets: increasing the levels of lipids with protective pathways (CE 18:3, CE 22:6, TG 56:5) and decreasing the levels of lipids with deleterious pathways (TG 51:3, TG 54:2, TG 56:4).

Our analysis showed that lipids within the same super class can significantly mediate the effects of *APOE2* on cognitive function in different directions. For example, we identified two glycerolipids with opposite effects. Increasing levels of TG 56:5 was protective for total-time and ink-time in both primary and sensitivity analysis. In contrast, increasing levels of TG 56:4 was deleterious for ink-time in both primary and sensitivity analysis. In addition, compared to the main analysis, the sensitivity analysis revealed more TGs with a statistically significant deleterious indirect effect (TG 51:3 and TG 54:2). The selective effects of these glycerolipids on total-time and ink-time of the CDT but not on think-time, suggest that they mediate graphomotor function components of CDT test performance more specifically. Furthermore, our findings of different TGs can help clarify their complex mechanisms on cognitive function and cognitive test performance, as such conflicting associations were also reported in previous studies [31–33].

The sterol lipids included in this analysis consistently showed their protective mediation of the effect of *APOE2*. CE 18:3 was protective for both total-time and think-time in the primary analysis and protective for think-time in the sensitivity analysis, and CE 22:6 was protective for think-time in the sensitivity analysis. This is aligned with recent research reporting that cholesterols are either not associated with or may even protect against late-life cognitive decline [34–36]. The relationship of the sterol lipids with total time and think time, rather than ink time, also suggests that they mediate cognitive processing components of CDT test performance more specifically.

Previous studies on mediation analysis of *APOE* have focused on mediators of cerebral blood flow [37], brain tissue volume [38], and neuropathological pathways [39], suggesting these factors partially mediated the negative effect of *APOE4*. Considering lipids as mediators, one study found that *APOE2* had a significant indirect negative effect on cognition through total cholesterol [15]. Another study found no lipids but BMI significantly mediated the risk of AD [16]. However, one study on the risk of AD found 11 lipid species that mediated the effect of *APOE2*, accounting for up to 30% of the total effect of *APOE2* on AD resilience [14]. This is aligned with our finding of 10%-27% mediated proportions from all lipid pathways. Compared to previous research, the novelty of our research lies in revealing both protective and deleterious pathways mediated through lipids as well as differential effects of glycerolipids and sterol lipids on graphomotor function and cognitive processing, respectively.

### Limitations

In our primary analysis, the confidence intervals for some lipids were marginally close to zero. For example, the lower limit of 95% CI for TG 51:3 was below zero for all CDT times in the primary analysis. The number of LLFS participants who completed the CDT are not large, which may have led to limited statistical power in those findings. Future research may aim to combine multiple studies with digital CDT and lipids data for an increased statistical power to detect more significant indirect effects through lipids.

Other limitations in this study include that we do not consider the potential interaction effect between *APOE* and lipids metabolites. Also, since our exposure, *APOE*, is determined at birth, all confounders in our analysis may potentially act as post-treatment confounders, complicating the causal interpretation. Additionally, all outcomes in this application focus on the CDT time outcomes, which may only reflect certain aspects of cognitive function, for example, processing speed. Our future research also plans to investigate the effects of *APOE* alleles and lipids on different domains of cognitive function using additional measures.

### Conclusions

We analyzed data from the Long Life Family Study to elucidate the relationship between *APOE* variants, lipids, and cognitive function measured by CDT times. The results revealed a direct protective effect of *APOE2* on cognitive and motor function and highlighted indirect effects through several lipid species that mediate the effects of *APOE2* in either a protective or a deleterious pathway. The results also showed that the combined indirect effect through all lipids can mediate 10%-27% of total effect of *APOE2* on cognitive function. Additionally, the identified protective and deleterious lipid pathways present potential opportunities for developing new therapeutics targeting these lipids to modulate the effects of *APOE2* on cognitive function.

## Data Availability

All data produced in the present study are available upon reasonable request to the authors.

## Acknowledgement

The authors would like to thank all LLFS participants for the generous donation of their time, biospecimens, and study data.

