## Supplementary files for "Causal mediation analysis of the neuroprotection of *APOE2* through lipid pathways"

### Supplementary File

##### Calculation of direct effects and indirect effects

We provide a detailed description of the computation of direct effects and indirect effects, following the methodology of VanderWeele (2014). We performed a regression approach to estimate the total, direct, and indirect effect of *APOE* on each of the outcomes, total-, ink-, and think-time of the CDT.

Step 1: we fitted the mediator regression models, one for each lipid, denoted as $M^{(i)}$

$$E\left[ log(M^{\left( i \right)}) | a,C \right]=\theta_{0}^{(i)}+\theta_{1}^{(i)}\cdot a_{APOE}+\boldsymbol{\theta}_{\boldsymbol{2}}^{\left( i \right)}\cdot\boldsymbol{C,}$$

where $M^{(i)}$ is the $i$th mediator (lipid), $i=1,2,\ldots,K$*,* $\theta_{0}^{(i)}$ is the intercept, $a_{APOE}$ is the *APOE* genotype group coded as *APOE3*=0, *APOE2*=1 and $\theta_{1}^{(i)}$ is the corresponding coefficient, $\boldsymbol{C}$ is a vector of baseline confounders (sex, education, age at enrollment, and indicator of lipid-lowering medication usage, and indicator of young/old generation) and $\boldsymbol{\theta}_{\boldsymbol{2}}^{\left( i \right)}$ is the vector of their coefficients.

Step 2: we fit the outcome regression model for each outcome, incorporating all lipids simultaneously:

$$E\left[ Y | a,M,C \right]=\beta_{0}+\beta_{1}\cdot a_{APOE}+\beta_{2}^{\left( 1 \right)}\cdot{log(M}^{\left( 1 \right)})+\ldots\beta_{2}^{\left( K \right)}\cdot log(M^{\left( K \right)})+\boldsymbol{\beta}\cdot\boldsymbol{C,}$$

where $Y$ is one of the three CDT times, $\beta_{0}$ is the intercept, $\beta_{1}$, $\beta_{2}^{i}$ and $\boldsymbol{\beta}$ are the corresponding coefficients.

- The direct effect of *APOE2* on the outcome is the estimate of $\beta_{1}$.
- The indirect effect of *APOE2* through the pathway of $i$th lipid metabolite is estimated as the product of the estimates $\hat{\theta}_{1}^{(i)}\cdot\hat{\beta}_{2}^{(i)}.$
- The combined indirect effect of all the lipid metabolites is estimated as $\sum_{i=1}^{K} \hat{\theta}_{1}^{(i)}\cdot\hat{\beta}_{2}^{(i)}$.
- The total effect is the sum of the direct effect and combined indirect effect: $\beta_{1}+\sum_{i=1}^{K} \hat{\theta}_{1}^{(i)}\cdot\hat{\beta}_{2}^{(i)}$.

To assess the statistical significance of the estimated direct and indirect effects, we conducted 1000 bootstrap resamples to construct confidence intervals.

##### Supplementary Figures

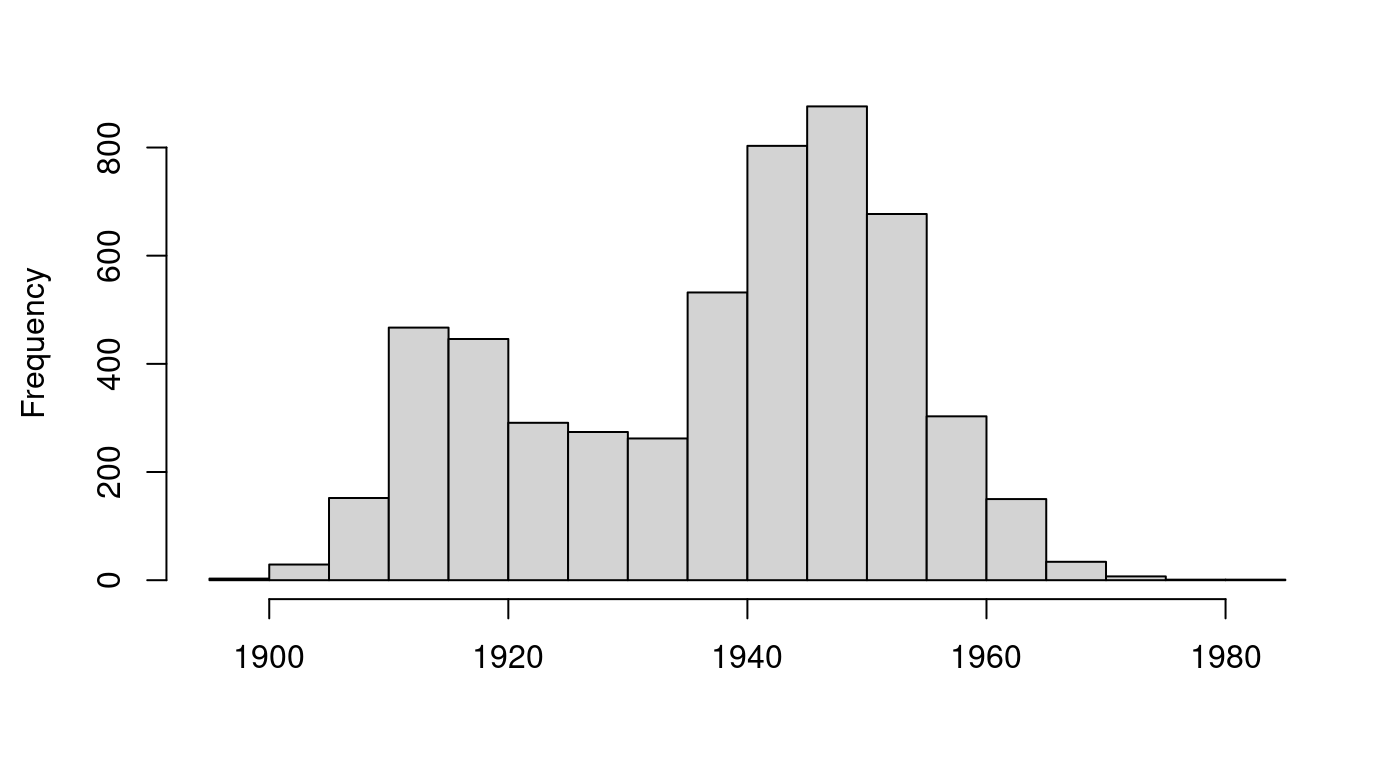

**Supplementary Figure 1**: Date of birth of the LLFS participants. The young/old generation is separated by whether birth year is >= 1935.

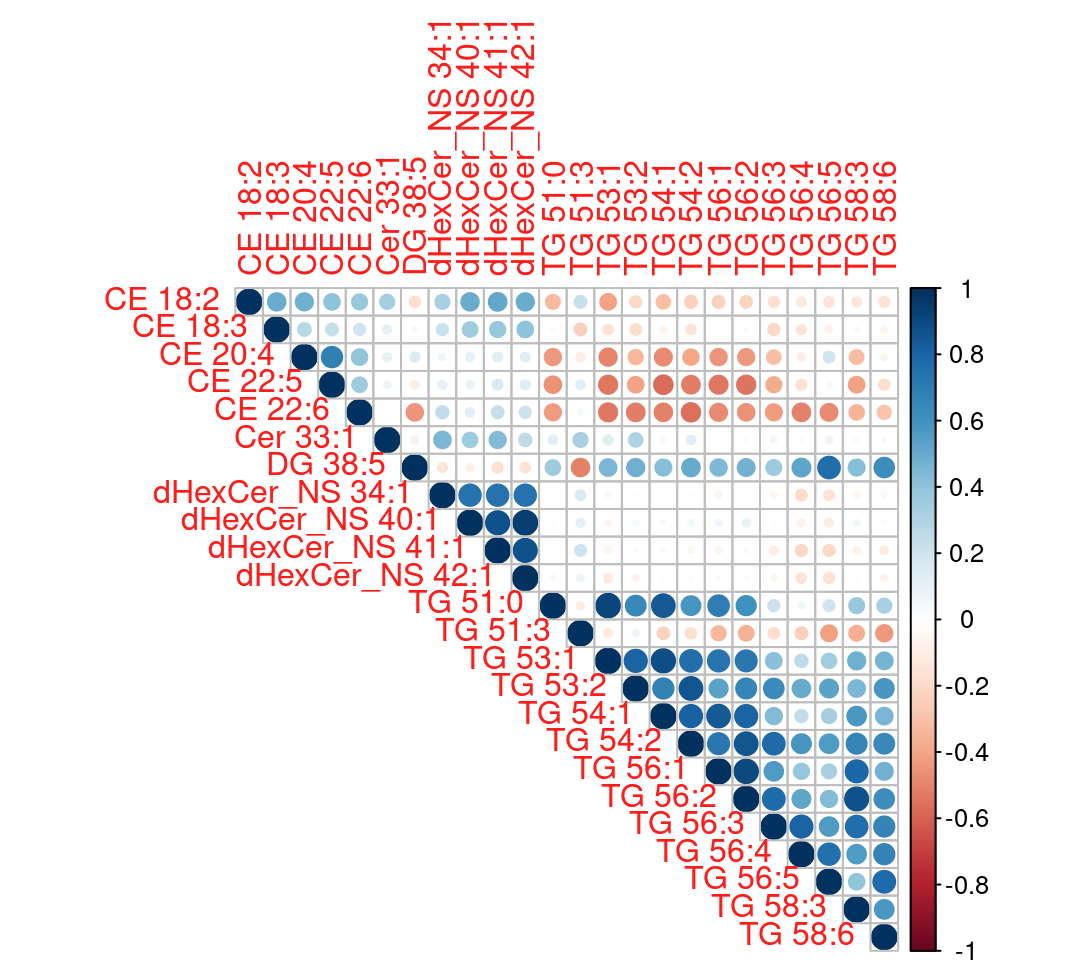

**Supplementary Figure 2**: The correlation matrix of the 24 lipids that were found to be significantly associated with *APOE* e2. We focused on the lipid pairs that has a correlation greater than 0.85: dHexCer_NS 40:1­­~ dHexCer_NS 41:1, dHexCer_NS 40:1­­~ dHexCer_NS 42:1, dHexCer_NS 41:1~ dHexCer_NS 42:1, TG 53:1 ~ TG 51:0, TG 53:1 ~ TG 54:1, TG 53:2 ~ TG 54:2, TG 54:2 ~ TG 56:2, TG 56:1 ~ TG 56:2, TG 56:2 ~ TG 58:3. There were multiple pairs involved common lipids, and five lipids are excluded from the model: dHexCer_NS 42:1, dHexCer_NS 40:1, TG 53:2, TG 54:1, TG 56:2.

##### Supplementary Tables: sensitivity analysis of digital CDT

| Total-time | | | Think-time | | | Ink-time | | |
| --- | --- | --- | --- | --- | --- | --- | --- | --- |
| Lipids | $\beta$ of *APOE3 vs APOE2* (SD) | P-value | Lipids | $\beta$ of *APOE3 vs APOE2* (SD) | P-value | Lipids | $\beta$ of *APOE3 vs APOE2* (SD) | P-value |
| CE 18:3 | -0.20 (0.06) | <0.01* | CE 18:3 | -0.20 (0.06) | <0.01* | DG 38:5 | 0.37 (0.06) | <0.01* |
| CE 22:6 | -0.22 (0.07) | <0.01* | CE 22:6 | -0.22 (0.07) | <0.01* | TG 51:3 | -0.29 (0.07) | <0.01* |
| Cer 33:1 | -0.16 (0.07) | 0.01* | Cer 33:1 | -0.16 (0.07) | 0.01* | TG 56:4 | 0.29 (0.07) | <0.01* |
| DG 38:5 | 0.37 (0.06) | <0.01* | DG 38:5 | 0.37 (0.06) | <0.01* | TG 56:5 | 0.29 (0.07) | <0.01* |
| dHexCer_NS 41:1 | -0.09 (0.06) | 0.13 | dHexCer_NS 41:1 | -0.09 (0.06) | 0.13 |  |  |  |
| TG 51:3 | -0.29 (0.07) | <0.01* | TG 51:3 | -0.29 (0.07) | <0.01* |  |  |  |
| TG 54:2 | 0.26 (0.07) | <0.01* | TG 54:2 | 0.26 (0.07) | <0.01* |  |  |  |
| TG 56:1 | 0.38 (0.08) | <0.01* | TG 56:1 | 0.38 (0.08) | <0.01* |  |  |  |
| TG 56:4 | 0.29 (0.07) | <0.01* | TG 56:3 | 0.36 (0.07) | <0.01* |  |  |  |
| TG 56:5 | 0.29 (0.07) | <0.01* | TG 56:5 | 0.29 (0.07) | <0.01* |  |  |  |

**Supplementary Table 1**: Results of the mediator regression in the sensitivity analysis on digital CDT times. After variable selection in the sensitivity analysis, less lipids stay in the model. Dependent variable: lipids (log scale and standardized). Independent variables: *APOE3* (reference group) vs *APOE2,* sex, education, age at enrollment, lipid-lowering medication usage, and indicator for young/old generation. $\beta$: estimated coefficients of E2 versus E3 (reference) on each lipid outcome. SD: standard deviation. *: p-value that reaches the significance level of 0.05.

| Total-time | | | Think-time | | | Ink-time | | |
| --- | --- | --- | --- | --- | --- | --- | --- | --- |
| Variables | $\beta$ (SD) | P-value | Variables | $\beta$ (SD) | P-value | Variables | $\beta$ (SD) | P-value |
| *APOE2* | **-3.02 (0.96)** | **<0.01** | *APOE2* | **-1.99 (0.70)** | **<0.01** | ***APOE2*** | **-1.16 (0.38)** | **<0.01** |
| CE 18:3 | 1.10 (0.56) | 0.05 | CE 18:3 | **1.09 (0.44)** | **0.01** | DG 38:5 | -0.49 (0.28) | 0.09 |
| CE 22:6 | 1.29 (0.79) | 0.10 | CE 22:6 | 1.09 (0.57) | 0.06 | **TG 51:3** | **-0.60 (0.24)** | **0.01** |
| Cer 33:1 | 0.95 (0.56) | 0.09 | Cer 33:1 | 0.75 (0.44) | 0.09 | **TG 56:4** | **0.77 (0.29)** | **0.01** |
| DG 38:5 | -1.61 (0.99) | 0.10 | DG 38:5 | -1.11 (0.74) | 0.13 | **TG 56:5** | **-1.01 (0.40)** | **0.01** |
| dHexCer_NS 41:1 | -0.94 (0.63) | 0.13 | dHexCer_NS 41:1 | -0.81 (0.47) | 0.08 |  |  |  |
| TG 51:3 | **-1.86 (0.78)** | **0.02** | TG 51:3 | -1.1 (0.58) | 0.06 |  |  |  |
| TG 54:2 | **2.86 (0.91)** | **<0.01** | TG 54:2 | **2.18 (0.72)** | **<0.01** |  |  |  |
| TG 56:1 | -1.27 (0.92) | 0.17 | TG 56:1 | -1.04 (0.70) | 0.14 |  |  |  |
| TG 56:4 | 1.77 (0.97) | 0.07 | TG 56:3 | 1.05 (0.80) | 0.19 |  |  |  |
| TG 56:5 | **-3.82 (1.45)** | **0.01** | TG 56:5 | **-2.33 (1.01)** | **0.02** |  |  |  |

**Supplementary Table 2**: Results of the outcome regression in the sensitivity analysis on digital CDT times. After variable selection in the sensitivity analysis, less lipids stay in the model. Dependent variable: CDT total-time, think-time, and ink-time. Independent variables: *APOE3* (reference group) vs *APOE2*, lipids (log scale and standardized), age at enrollment, sex, education, lipid-lowering medication usage, and indicator of young/old generation. $\beta$: estimated coefficients. SD: standard deviation. Bold font indicates the estimated coefficient reaching the significance level of 0.05.

| Total time | | Think time | | Ink time | |
| --- | --- | --- | --- | --- | --- |
| Lipids | Effect estimate (95% CI) | Lipids | Effect estimate (95% CI) | Lipids | Effect estimate (95% CI) |
| CE 18:3 | -0.22 (-0.52, 0.02) | CE 18:3 | **-0.22 (-0.49, -0.02)** | DG 38:5 | -0.18 (-0.45, 0.05) |
| CE 22:6 | -0.29 (-0.72, 0.1) | CE 22:6 | **-0.24 (-0.56, -0.01)** | TG 51:3 | **0.17 (0.04, 0.39)** |
| Cer 33:1 | -0.16 (-0.48, 0.02) | Cer 33:1 | -0.12 (-0.39, 0.03) | TG 56:4 | **0.23 (0.05, 0.47)** |
| DG 38:5 | -0.59 (-1.4, 0.2) | DG 38:5 | -0.41 (-1.02, 0.15) | TG 56:5 | **-0.29 (-0.61, -0.06)** |
| dHexCer_NS 41:1 | 0.09 (-0.07, 0.43) | dHexCer_NS 41:1 | 0.07 (-0.03, 0.32) |  |  |
| TG 51:3 | **0.54 (0.07, 1.27)** | TG 51:3 | 0.32 (-0.01, 0.82) |  |  |
| TG 54:2 | **0.74 (0.22, 1.47)** | TG 54:2 | **0.56 (0.17, 1.21)** |  |  |
| TG 56:1 | -0.48 (-1.39, 0.19) | TG 56:1 | -0.39 (-1.04, 0.18) |  |  |
| TG 56:4 | 0.52 (-0.02, 1.25) | TG 56:3 | 0.38 (-0.22, 1.13) |  |  |
| TG 56:5 | **-1.09 (-2.31, -0.31)** | TG 56:5 | **-0.67 (-1.52, -0.11)** |  |  |

**Supplementary Table 3**: Results of indirect effects of *APOE2* on CDT times through each lipid pathway after variable selection. 95% confidence intervals (CI) are generated using bootstrap percentile method. Protective lipids mediated the effect of *APOE2* to reduce CDT times, while deleterious lipids mediated the effect of *APOE2* to increase CDT times. Bold font indicates the effect estimates reaching the significance level of 0.05.
